# High Prevalence of Group A Rotavirus Among Children Under Five with Acute Diarrhea in Mogadishu: Unexpected Risk and Protective Factors, 2025

**DOI:** 10.1101/2025.11.19.25340609

**Authors:** Mohamed Yusuf Abdi, Deka Adan Hassan, Maaria Sacdi Mohamud

## Abstract

**Introduction:** Group A *rotavirus* is the leading cause of severe diarrhea in children under five years of age. This study aimed to evaluate the prevalence and risk factors associated with Group A *rotavirus* infection among children under five years of age with acute diarrhea in Mogadishu, Somalia.

**Methods:** A hospital-based cross-sectional study was conducted at two major hospitals in Mogadishu between February and March 2025. Children under five years of age with acute watery diarrhea were enrolled. A structured questionnaire was used to collect demographic data and risk factors from guardians. Stool specimens were analyzed for *rotavirus* antigens using an enzyme-linked immunosorbent assay. Bivariate analysis was performed using the chi-square test, and variables with *p* < 0.20 were entered into multivariable logistic regression to identify independent predictors of infection.

**Results:** Among 279 children under five with diarrhea, 113 (40.5%) tested positive for *rotavirus*. In multivariable analysis, tap water was significantly protective (AOR = 0.479, p = 0.006), while handwashing was unexpectedly linked to increased risk (AOR = 2.439, p = 0.031). Other factors were not significantly associated with infection.

**Conclusion:** The high prevalence of *rotavirus* and the protective link with tap water use highlight the need to improve water access and hygiene. Further investigation into hygiene practices is warranted to clarify the association with handwashing.

## Introduction

Diarrhea is the third leading cause of death among children under five globally, accounting for approximately 443,832 deaths annually, mostly in South Asia and sub-Saharan Africa [1]. *Rotavirus* continues to be a major etiologic agent of severe and fatal diarrhea in this age group, with especially high mortality in sub-Saharan Africa [2]. Of the seven *rotavirus* groups (A–G), Group A accounts for more than 90% of human infections [3]. *Rotavirus* gastroenteritis causes fever, vomiting, and watery diarrhea; severe cases may lead to dehydration, seizures, or death [4].

Although recent global estimates are limited, available data indicate that *rotavirus* remains a major public health threat, especially in low- and middle-income countries. In 2021, *rotavirus* was estimated to cause approximately 108,322 deaths, representing 24.4% of all diarrheal deaths among children under five worldwide [2]. A meta-analysis of studies from 2009 to 2022 reported that the pooled prevalence of *rotavirus* infection among African children with gastroenteritis was 30.8% (95% CI: 28.1–31.5%), with a case fatality rate of 1.2% (95% CI: 0.7–2.0%) [5]. The high mortality burden in Africa underscores the urgent need for widespread *rotavirus* vaccination, improved hygiene, and strengthened surveillance systems.

The WHO recommends *rotavirus* vaccination in all national programs. Four oral vaccines, Rotarix, rotateq, Rotavac, and rotasiil, are WHO-prequalified and effective against severe gastrointestinal illness [6]. By 2023, 38 of 47 countries in the WHO African Region had introduced *rotavirus* vaccination into their national immunization programs [7]. Building on this progress, Somalia, with support from Gavi, WHO, and UNICEF, officially introduced the *rotavirus* vaccine to its national program in April 2025 [8–10]. Prior to the introduction of the *rotavirus* vaccine, *rotavirus* caused 41.4% of childhood diarrhea hospitalizations in (2023) and 33.5% of cases in Mogadishu in (2024) [11,12].

While earlier research examined *rotavirus* infections among children in Mogadishu, Somalia, there remains a need for updated, targeted studies using more sensitive and specific detection methods to better understand the current contributing factors. Therefore, this study aimed (1) to determine the prevalence of Group A *rotavirus* infection among children under five years of age with acute diarrhea in Mogadishu, Somalia, and (2) to identify factors associated with *rotavirus* infection using enzyme-linked immunosorbent assay (ELISA) for diagnosis.

## Methods

### Study design

A hospital-based cross-sectional study was conducted to determine the prevalence of Group A *rotavirus* infection and associated factors among children under five years with acute diarrhea in Mogadishu, Somalia.

### Study Setting and Population

The study was conducted at Royal Hospital (Hodan district) and SOS Hospital (Huriwaa district), both in Mogadishu, Somalia, between February 1 and March 31, 2025. Mogadishu is the capital city of Somalia, located in the Benadir Region along the Indian Ocean coast (latitudes 2°00′–2°10′ N, longitudes 45°15′–45°25′ E; altitude ∼9 m). Children aged 0–5 years with acute, non-bloody diarrhea were eligible. Children with hospital-acquired gastroenteritis, prior antimicrobial use, or whose caregivers refused consent were excluded.

### Variables

#### Outcome variable

*Rotavirus* infection, defined as a positive VP6 antigen result by ELISA.

#### Predictor variables

Child-related factors (age, sex, breastfeeding, nutritional status, kindergarten attendance, diarrhea severity), maternal and household factors (age, education, occupation, marital status, family size, presence of other children under five), and environmental/behavioral factors (handwashing practices, access to safe drinking water, number of household rooms).

### Data Sources and Measurement

A structured, interview-based questionnaire as developed from a literature review [13–17], adapted for cultural and contextual relevance and pre-tested on 5% of the study population. Three public health specialists assessed content validity. The interviews were conducted face-to-face with children’s mothers to gather requisite information by trained health professionals. The questionnaire included sections on child-related factors, maternal and household factors, and behavioral and environmental factors. Clinical data were extracted from hospital medical records. Mothers were given sterile stool containers for their children, and samples were transported on ice to the Microbiology Lab at SIMAD University. *Rotavirus* VP6 antigen detection was performed using a commercial ELISA kit (prospect *Rotavirus* Microplate Kit, Oxoid Ltd., UK) according to manufacturer instructions.

### Sample size

A sample of 279 children was calculated using a single-population proportion formula based on a 23.8% prevalence reported in a previous study [18], 95% confidence level, and 5% margin of error. Using simple random sampling, 56% were enrolled from Royal Hospital and 44% from SOS Hospital.

### Data analysis

Data was cleaned and analyzed using the Statistical Package for Social Sciences version 27. Categorical variables were summarized using frequencies and proportions. To determine the factors associated with *rotavirus* diarrhea, binary logistic regression was used to calculate CORS with 95% CIs; p < 0.05 indicated significance. Variables with p < 0.2 in bivariate analysis were included in the multivariate model to control for confounding. AORS with 95% CIs were reported, with p < 0.05 considered significant. Missing data was minimal and excluded listwise.

### Ethical consideration

This study received ethical approval from the SIMAD University Institutional Review Board (Ref: 2025/SU-IRB/FMHS/P0028) on 17 January 2025, with additional authorization from both participating study sites, Royal Hospital and SOS Hospital. Data collection was conducted from 1 February 2025 to 31 March 2025. Written informed consent was obtained from caregivers prior to stool sample collection and completion of questionnaires. Caregivers were informed about the study purpose, procedures, potential risks, and their rights, including the right to withdraw at any time without affecting the child’s care. Clinical data were also extracted from existing hospital medical records. The ethics committee granted a waiver of the requirement for additional consent for the use of these records. The authors had no access to identifying information during or after data collection. All collected data are secured in password-encrypted files accessible only to the researchers. Electronic data were stored on a password-protected computer accessible only to the research team.

## Results

### Sociodemographic and clinical characteristics of children

A total of 279 children under five years were included in the study, and stool samples were successfully tested for *rotavirus* in all participants. No demographic, clinical, or household data were missing. Of these children, 50.9% male and 49.1% female. Most were aged 1–11 months (45.5%) or 25–59 months (48.0%), with 6.5% aged 12–24 months. Breastfeeding was reported in 62.0% of cases. About 81.7% were well nourished, while 18.3% were malnourished. Only 8.6% attended kindergarten. Illness severity was mild in 40.9%, moderate in 22.9%, and severe in 36.2% “Table 1”.

**Table 1.** Sociodemographic and clinical characteristics of children under five with acute diarrhea, recruited from Royal and SOS Hospitals in Mogadishu, Somalia, in 2025 (N=279)

| Variable | Category | Frequency (%) |
| --- | --- | --- |
| Gender | Male | 142 (50.9%) |
|  | Female | 137 (49.1%) |
| Age | 1–11 months | 127 (45.5%) |
|  | 12–24 months | 18 (6.5%) |
|  | 25–59 months | 134 (48.0%) |
| Brestfeeding | Yes | 173 (62.0%) |
|  | No | 106 (38.0%) |
| Nutritional status | Well-nourished | 228 (81.7%) |
|  | Malnutrition | 51 (18.3%) |
| Kindergarten attending | Yes | 24 (8.6%) |
|  | No | 255 (91.4%) |
| **Severity of illness** | Mild | 114 (40.9%) |
|  | Moderate | 64 (22.9%) |
|  | Severe | 101 (36.2%) |

### Descriptive profile of mothers and households

Most mothers were aged 25–34 (67.0%), with 42.7% having no formal education and 35.1% holding a diploma or higher. About 42.3% were employed, and 64.9% were married. Most households (66.7%) had over five members, and 89.6% had one or two children under five. Hand washing was reported by 88.5% of mothers. Commercial water was the main source (53.8%), and 86.4% lived in homes with three or more rooms “Table 2”.

**Table 2.** Maternal and household characteristics of children under five with acute diarrhea, recruited from Royal and SOS Hospitals in Mogadishu, Somalia, in 2025 (N=279).

| Variable | Category | Frequency (%) |
| --- | --- | --- |
| **Mother's age (years)** | 18- 24 years | 43 (15.4%) |
|  | 25- 34 years | 187 (67.0%) |
|  | >35 | 49 (17.6%) |
| **Mother educational level** | Illiterate or No Formal education | 119 (42.7%) |
|  | Primary/Secondary | 62 (22.2%) |
|  | Diploma and above | 98 (35.1%) |
| **Occupational status** | Employed | 118 (42.3%) |
|  | Unemployed/student | 161 (57.7%) |
| **Marital status** | Married | 181 (64.9%) |
|  | Widowed/divorced | 98 (35.1%) |
| **Family size** | ≤3 | 4 (1.4%) |
|  | 4-5 | 89 (31.9%) |
|  | >5 | 186 (66.7%) |
| **Other Children less than 5 years** | ≤2 | 250 (89.6%) |
|  | ≥2 | 29 (10.4%) |
| **Hand washing** | Yes | 247 (88.5%) |
|  | No | 32 (11.5%) |
| **Source of drinking water** | Tap water | 129 (46.2%) |
|  | Commercial water | 150 (53.8%) |
| **Household's rooms numbers** | <3 | 38 (13.6%) |
|  | ≥3 | 241 (86.4%) |

### Prevalence of *rotavirus* infection

Out of 279 children tested, 113 (40.5%) were positive for *rotavirus* antigens, while 166 (59.5%) tested negative.

### Factors associated with *rotavirus* infection

**In bivariable analysis**, children of mothers aged 18–24 had significantly higher odds of *rotavirus* infection compared to those whose mothers were over 35 (OR = 2.619, 95% CI: 1.107–6.197, p = 0.028). Tap water use was associated with reduced odds of infection (OR = 0.537, 95% CI: 0.330–0.875, p = 0.013). Other factors, including marital status and handwashing, were not statistically significant “Table 3”.

**Table 3.**
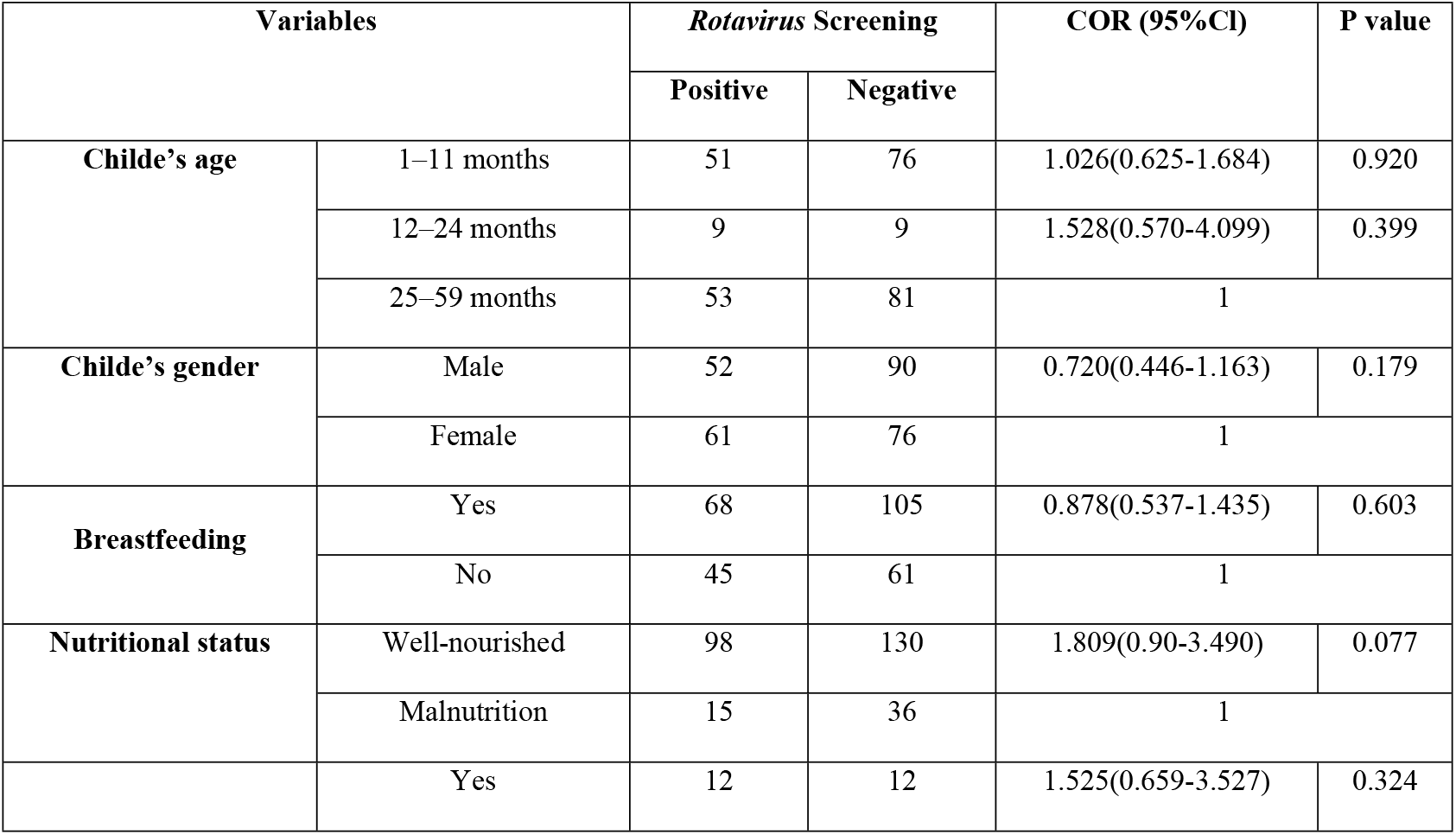
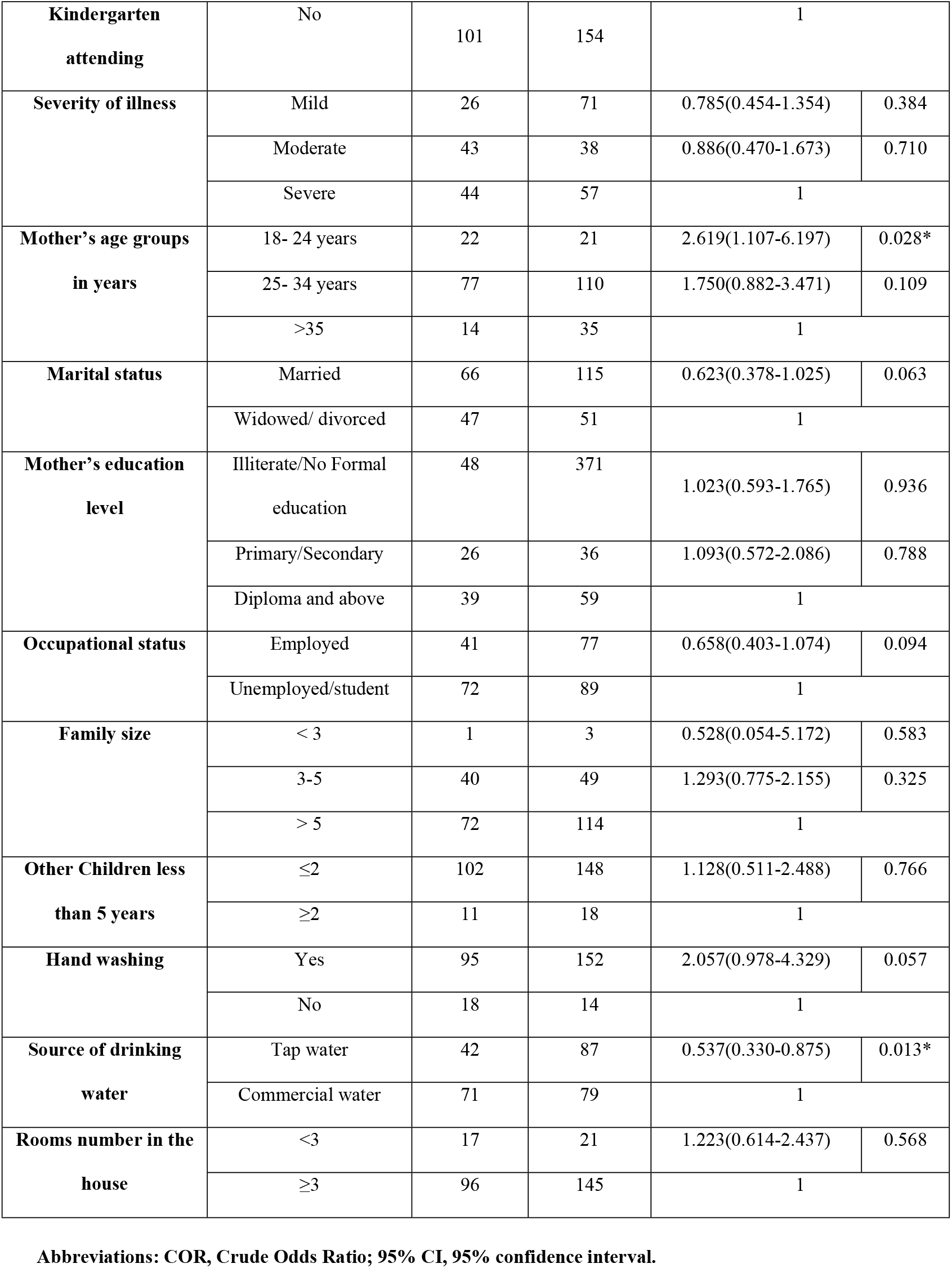
Bivariable logistic regression analysis of factors associated with rotavirus infection among children under five with acute diarrhea, recruited from Royal and SOS Hospitals in Mogadishu, Somalia, in 2025 (N=279).

**In multivariate analysis**, using tap water (AOR = 0.479, 95% CI: 0.28–0.82, p = 0.006) and having a married mother (AOR = 0.590, 95% CI: 0.34–1.01, p = 0.053) were protective against *rotavirus* infection. Male children also had lower odds (AOR = 0.749, 95% CI: 0.45–1.24, p = 0.265), though not statistically significant. Handwashing (AOR = 2.439, 95% CI: 1.09–5.45, p = 0.031) and good nutritional status (AOR = 1.893, 95% CI: 0.94–3.80, p = 0.074) were associated with higher odds. Other factors assessed in the model did not show significant associations “Table 4”.

**Table 4.**
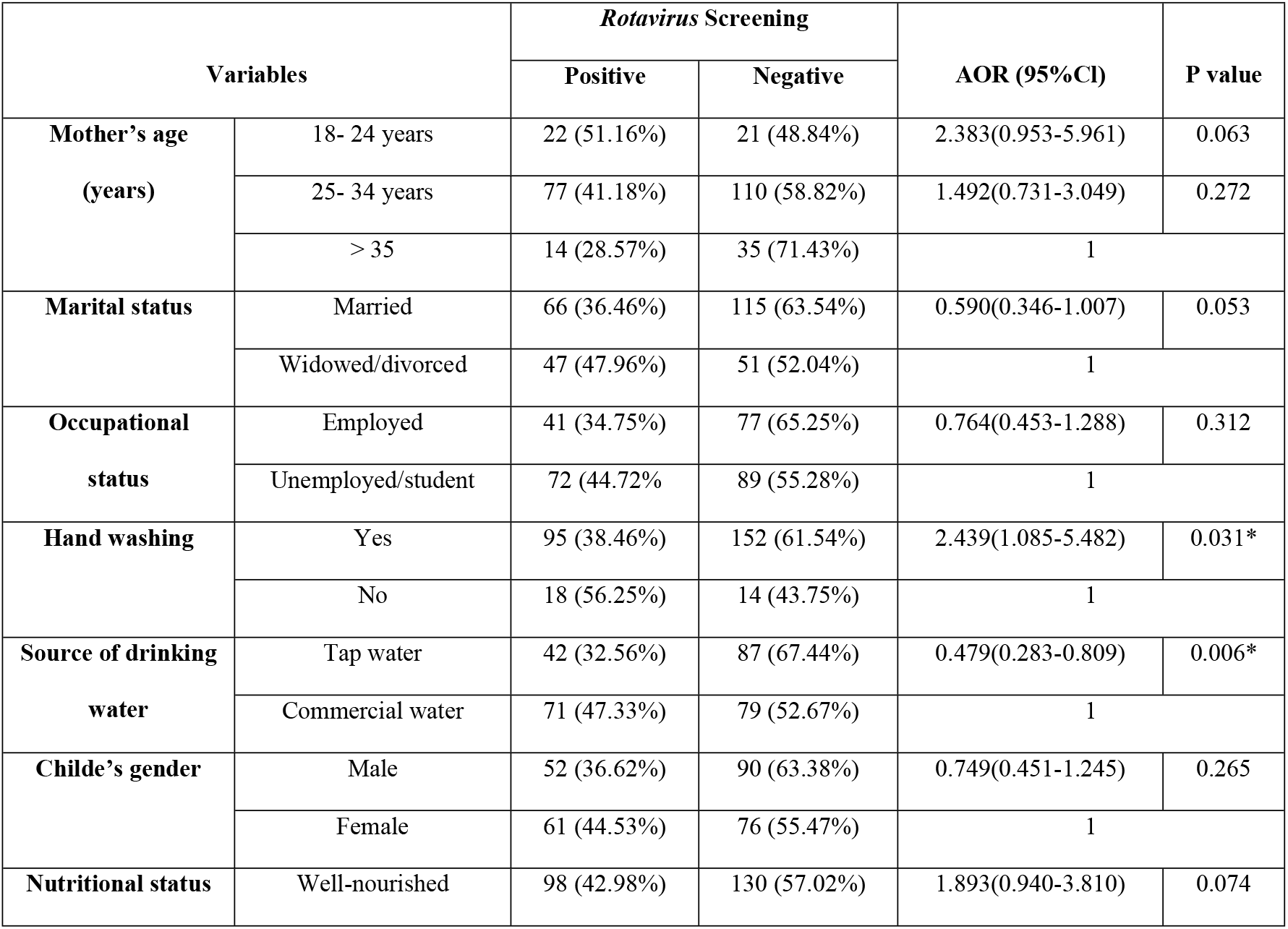
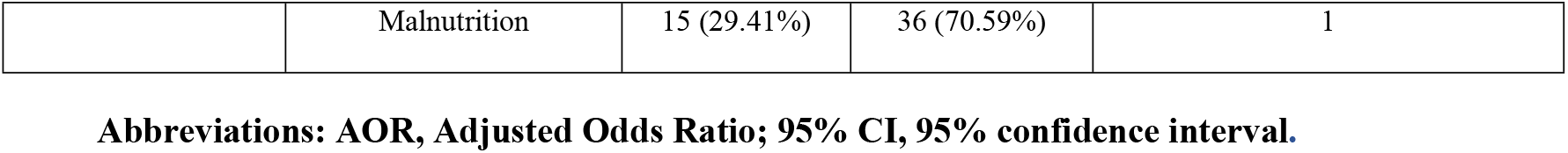
Multivariate logistic regression analysis of factors associated with *rotavirus* infection among children under five with acute diarrhea, recruited from Royal and SOS Hospitals in Mogadishu, Somalia, in 2025 (N=279).

| Variables |  | Rotavirus Screening |  | AOR (95%CI) | P value |
| --- | --- | --- | --- | --- | --- |
|  |  | Positive | Negative |  |  |
| Mother's age<br>(years) | 18- 24 years | 22 (51.16%) | 21 (48.84%) | 2.383(0.953-5.961) | 0.063 |
|  | 25- 34 years | 77 (41.18%) | 110 (58.82%) | 1.492(0.731-3.049) | 0.272 |
|  | > 35 | 14 (28.57%) | 35 (71.43%) | 1 |  |
| Marital status | Married | 66 (36.46%) | 115 (63.54%) | 0.590(0.346-1.007) | 0.053 |
|  | Widowed/divorced | 47 (47.96%) | 51 (52.04%) | 1 |  |
| Occupational<br>status | Employed | 41 (34.75%) | 77 (65.25%) | 0.764(0.453-1.288) | 0.312 |
|  | Unemployed/student | 72 (44.72%) | 89 (55.28%) | 1 |  |
| Hand washing | Yes | 95 (38.46%) | 152 (61.54%) | 2.439(1.085-5.482) | 0.031* |
|  | No | 18 (56.25%) | 14 (43.75%) | 1 |  |
| Source of drinking<br>water | Tap water | 42 (32.56%) | 87 (67.44%) | 0.479(0.283-0.809) | 0.006* |
|  | Commercial water | 71 (47.33%) | 79 (52.67%) | 1 |  |
| Childe's gender | Male | 52 (36.62%) | 90 (63.38%) | 0.749(0.451-1.245) | 0.265 |
|  | Female | 61 (44.53%) | 76 (55.47%) | 1 |  |
| Nutritional status | Well-nourished | 98 (42.98%) | 130 (57.02%) | 1.893(0.940-3.810) | 0.074 |

|  |  |  |  |  |
| --- | --- | --- | --- | --- |
|  | Malnutrition | 15 (29.41%) | 36 (70.59%) | 1 |

**Abbreviations: AOR, Adjusted Odds Ratio; 95% CI, 95% confidence interval.**

## Discussion

This study revealed that tap water use was protective against *rotavirus* infection, whereas handwashing was associated with increased odds of infection. The prevalence of *rotavirus* among children under five with diarrhea was 40.5%, which was comparable to a previous local study (41.4%) [11], slightly higher than a more recent one (33.5%) [13], and consistent with findings from Southern Karnataka, India (38.3%) [19]. Nevertheless, this figure was lower than those reported in other studies from Chad (51.81%) [20], Cameroon (54.6%) [21], and Vietnam (68.8%) [22], but higher than in Chad (12.76%) [23]; Kenya (15.2%) [24]; Tanzania (26.4%) [25]; Nigeria (31.5%) [26]; Iran (36.6%) [27]; India (35.5%) [28]; and Democratic Republic of the Congo (DRC) (36%) [15]. Prevalence variation may reflect climate, environment, socioeconomic, cultural, diagnostic, sampling, and vaccination differences.

In this study, *rotavirus* peaked at 12–24 months (50%) but age was not a significant risk factor. Our findings match a Nepal study showing higher infection under 24 months, though it found age significant [29]. Similar results were also documented in Uganda [16,30]. This difference may stem from sample size, population traits, or exposure and immunity variations.

Consistent with previous studies [16,22,26,30–34], this study found no significant links between *rotavirus* infection and child factors like gender, breastfeeding, nutrition, or illness severity. Males had lower infection odds than females, and well-nourished children had higher odds than malnourished, but neither was statistically significant. Unlike this, two Ugandan studies found significant links between male sex, breastfeeding, and *rotavirus* diarrhea [16,30].

This variation may result from factors like exclusive breastfeeding, maternal immunity, or hygiene during feeding.

Maternal age’s link to *rotavirus* infection is complex: bivariable analysis showed lower odds for children of mothers 18–24 versus over 35, but multivariable analysis showed higher, non-significant odds. This aligns with studies from China, Uganda, and Vietnam, which also found no significant association between maternal age and *rotavirus* infection [16,22,33].

Marital status showed a significant association in bivariable and marginally in multivariable analysis; children of married mothers had lower infection odds than those of widowed or divorced mothers. This suggests family support may improve child health through better care and stability. However, other studies from Eritrea, DRC, and Tanzania reported no significant associations, possibly due to cultural, social, and healthcare differences [15,31,34].

An unexpected finding was that handwashing lowered infection odds in bivariable analysis but showed higher odds in multivariable analysis. Still, this supports evidence that hand hygiene helps prevent fecal–oral transmission, especially of *rotavirus*. This aligns with studies from Ethiopia showing that children of caregivers who practiced handwashing had significantly lower *rotavirus* risk [35]. In contrast, a study from Indonesia found no significant link between handwashing and *rotavirus* infection [36]. This difference might be due to variations in cultural hygiene practices.

In both analyses, children using tap water had lower *rotavirus* risk than those using commercial water, suggesting treated tap water may offer better protection against contamination. These findings are consistent with studies from Uganda [16] and the DRC [15], which reported lower infection rates among children using tap water versus sources like well water, though significance varied. Conversely, a study from Tanzania [34] found no significant relationship, suggesting that local water quality, infrastructure, and hygiene behaviors may influence outcomes. The study’s hospital-based setting in Mogadishu limits the findings’ representativeness of all Somali children; yet, alignment with East African research suggests generalizability to similar urban, low-income settings.

The limitations of this study include that its cross-sectional design prevented causal inferences, caregiver-reported data may have introduced recall and social desirability bias, the short data collection period limited assessment of seasonal trends, and environmental factors were not fully evaluated.

## Conclusion

This study identified a high prevalence (40.5%) of Group A *rotavirus* among children under five with acute diarrhea in Mogadishu. Tap water use was significantly protective against infection, while handwashing was associated with higher odds of infection. Other factors were not significantly associated with *rotavirus* infection. These results underscore the importance of enhancing water quality, promoting better hygiene practices, and implementing targeted public health strategies.

## Data Availability

All data supporting the results of this study, including patient demographics, rotavirus test results, and questionnaire responses, are available in the supplementary files and from the corresponding author upon reasonable request.

## Acknowledgements

We sincerely thank the staff of the partnering healthcare institutions for their vital support and the research participants for their valuable contributions to this study.

## Author Contributions

All authors contributed significantly to the study’s design, data collection, analysis, and manuscript preparation. They meet the ICMJE authorship criteria, have approved the final version, and agree to its submission. All materials will be made available for non-commercial research use.

## References

1. World Health Organization. Diarrhoeal disease. 2024 [cited 2025 Aug 25]. Available from: https://www.who.int/news-room/fact-sheets/detail/diarrhoeal-disease.

2. Black RE, Perin J, Yeung D, Rajeev T, Miller J, Elwood SE, et al. Estimated global and regional causes of deaths from diarrhoea in children younger than 5 years during 2000–21: a systematic review and Bayesian multinomial analysis. Lancet Glob Health. 2024;12(6):e919– e928. doi: 10.1016/S2214-109X(24)00078-0.

3. Pan American Health Organization. Rotavirus. 2024 [cited 2025 Aug 25]. Available from: https://www.paho.org/en/topics/rotavirus.

4. Dian Z, Sun Y, Zhang G, Xu Y, Fan X, Yang X, et al. Rotavirus-related systemic diseases: clinical manifestation, evidence and pathogenesis. Crit Rev Microbiol. 2021;47(5):580–595. doi: 10.1080/1040841X.2021.1907738.

5. Njifon HLM, Kenmoe S, Ahmed SM, Roussel Takuissu G, Ebogo-Belobo JT, Njile DK, et al. Epidemiology of Rotavirus in Humans, Animals, and the Environment in Africa: A Systematic Review and Meta-analysis. J Infect Dis. 2024;229(5):1470–1480. doi: 10.1093/infdis/jiad500.

6. World Health Organization. Immunization, Vaccines and Biologicals, Rotavirus. 2024 [cited 2025 Aug 15]. Available from: https://www.who.int/teams/immunization-vaccines-and-biologicals/diseases/rotavirus.

7. Mwenda JM, Mandomando I, Worwui AK, Gacic-Dobo M, Katsande R, Bwaka AM, et al. A decade of rotavirus vaccination in the World Health Organization African Region: An in-depth analysis of vaccine coverage from 2012 to 2023. Vaccine. 2025;48:126768. doi: 10.1016/j.vaccine.2025.126768.

8. World Health Organization. Somalia launches lifesaving vaccines to prevent pneumonia and diarrhoea in children. 2025 [cited 2025 Aug 26]. Available from: https://www.emro.who.int/somalia/news/somalia-launches-lifesaving-vaccines-to-prevent-pneumonia-and-diarrhoea-in-children.html.

9. Gavi, the Vaccine Alliance. Federal Republic of Somalia launches nationwide vaccination against pneumococcal disease and rotavirus diarrhoea. 2025 [cited 2025 Aug 26]. Available from: https://www.gavi.org/news/media-room/somalia-launches-nationwide-vaccination-against-pneumococcal-disease-and-rotavirus.

10. UNICEF. Somalia launches two lifesaving vaccines to prevent pneumonia and diarrhea in children. 2025 [cited 2025 Aug 26]. Available from: https://www.unicef.org/somalia/press-releases/somalia-launches-two-lifesaving-vaccines.

11. Orhan Z, Mohamud S, Mohamud R, Doğan S. Rotavirus prevalence in children with acute gastroenteritis admitted to a Tertiary Hospital in Somalia in 2020-2023: a retrospective, single-center study. Public Health Manag Technol. 2024;15:365–373. doi: 10.2147/PHMT.S475345.

12. Roble MA, Anshur YAA, Ali AA, Ahmed FH, Adan AM, Ali NA, et al. Prevalence of Rotavirus Infection among Hospitalized Children Under Five Years of Age with Acute Diarrhea in Mogadishu, Somalia. Asian J Med Health. 2024;22(11):181–191. doi: 10.9734/ajmah/2024/v22i111129.

13. Chissaque A, Cassocera M, Gasparinho C, Langa JS, Bauhofer AFL, Chilaúle JJ, et al. Rotavirus A infection in children under five years old with a double health problem: undernutrition and diarrhoea – a cross-sectional study in four provinces of Mozambique. BMC Infect Dis. 2021;21(1):18. doi: 10.1186/s12879-020-05718-9.

14. Ali KB, Gadzama GB, Zailani SB, Mohammed Y, Daggash BB, Yakubu YM, et al. The Risk Factors Associated with Rotavirus Gastroenteritis among Children Under Five Years at University of Maiduguri Teaching Hospital, Borno State, Nigeria. Niger J Med. 2022;31(1):35–40. doi: 10.4103/NJM.NJM_114_21.

15. Gbebangi-Manzemu D, Kampunzu VM, Vanzwa HM, Mumbere M, Bukaka GM, Likele BB, et al. Clinical profile of children under 5 years of age with rotavirus diarrhoea in a hospital setting in Kisangani, DRC, after the introduction of the rotavirus vaccine, a cross-sectional study. BMC Pediatr. 2023;23(1):193. doi: 10.1186/s12887-023-04022-0.

16. Laker G, Nankunda J, Melvis BM, Kajoba D, Nduwimana M, Kimera J, et al. Prevalence and factors associated with rotavirus diarrhea among children aged 3–24 months after the introduction of the vaccine at a referral hospital in Uganda: a cross-sectional study. BMC Pediatr. 2024;24(1):358. doi: 10.1186/s12887-024-04842-8.

17. Getachew A, Tadie A, G. Hiwot M, Guadu T, Haile D, G. Cherkos T, et al. Environmental factors of diarrhea prevalence among under five children in rural area of North Gondar zone, Ethiopia. Ital J Pediatr. 2018;44(1):95. doi: 10.1186/s13052-018-0540-7.

18. Konstantopoulos A, Tragiannidis A, Fouzas S, Kavaliotis I, Tsiatsou O, Michailidou E, et al. Burden of rotavirus gastroenteritis in children <5 years of age in Greece: hospital-based prospective surveillance (2008–2010). BMJ Open. 2013;3(12):e003570. doi: 10.1136/bmjopen-2013-003570.

19. Shetty RS, Kamath VG, Nayak DM, Hegde A, Saluja T. Rotavirus associated acute gastroenteritis among under-five children admitted in two secondary care hospitals in southern Karnataka, India. Clin Epidemiol Glob Health. 2017;5(1):28–32. doi: 10.1016/j.cegh.2016.06.002.

20. Ndifor F, Lawane AI, Ngam-Asra N, Adoum MA, Otchom BB, Alio HM. The Prevalence of Rotavirus and Adenovirus in Children 0-5 years old Suffering from Acute Diarrhea in the University Hospital Center of Mother and Child (UHC-MC) in N’Djamena, Chad. Int J Sci Adv. 2021;2(6). doi: 10.51542/ijscia.v2i6.7.

21. Ghapoutsa RN, Boda M, Gautam R, Ndze VN, Mugyia AE, Etoa F-X, et al. Detection of diarrhoea associated rotavirus and co-infection with diarrhoeagenic pathogens in the Littoral region of Cameroon using ELISA, RT-PCR and Luminex xtag GPP assays. BMC Infect Dis. 2021;21(1):614. doi: 10.1186/s12879-021-06318-x.

22. Van Chuc D, Linh DP, Linh DV, Van Linh P. Clinical Epidemiology Features and Risk Factors for Acute Diarrhea Caused by Rotavirus A in Vietnamese Children. Int J Pediatr. 2023;2023:1–7. doi: 10.1155/2023/4628858.

23. Djikoloum B, Abakar MF, Ndze VN, Nkandi RG, Enjeh CN, Kimala P, et al. Epidemiology of group A rotavirus in children under five years of age with gastroenteritis in N’Djamena, Chad. BMC Infect Dis. 2024;24(1):111. doi: 10.1186/s12879-023-08647-5.

24. Gikonyo J, Mbatia B, Okanya P, Obiero G, Sang C, Nyangao J. Rotavirus prevalence and seasonal distribution post vaccine introduction in Nairobi county Kenya. Pan Afr Med J. 2019;33. doi: 10.11604/pamj.2019.33.269.18203.

25. Mchaile DN, Philemon RN, Kabika S, Albogast E, Morijo KJ, Kifaro E, et al. Prevalence and genotypes of Rotavirus among children under 5 years presenting with diarrhoea in Moshi, Tanzania: a hospital based cross sectional study. BMC Res Notes. 2017;10(1):542. doi: 10.1186/s13104-017-2883-3.

26. Ojobor CD, Olovo CV, Onah LO, Ike AC. Prevalence and associated factors to rotavirus infection in children less than 5 years in Enugu State, Nigeria. Virusdisease. 2020;31(3):316– 322. doi: 10.1007/s13337-020-00614-x.

27. Sedighi P, Karami M, Razzaghi M, Emamjamaat M, Karimi A, Mansour Ghanaiee R, et al. The Frequency of Rotavirus Gastroenteritis in Children from West of Iran and Genotyping of Rotavirus Isolates: A Suggestion for Further Changes in Childhood Immunization Program. J Res Health Sci. 2024;24(3):e00621. doi: 10.34172/jrhs.2024.156.

28. Giri S, Nair NP, Mathew A, Manohar B, Simon A, Singh T, et al. Rotavirus gastroenteritis in Indian children < 5 years hospitalized for diarrhoea, 2012 to 2016. BMC Public Health. 2019;19(1):69. doi: 10.1186/s12889-019-6406-0.

29. Shrestha J, Shrestha SK, Mason C, Sornsakrin S, Silapong S, Dhakwa J, et al. Rotavirus strains in children less than 5 years of age: A case control study. J Clin Virol Plus. 2024;4(2):100183. doi: 10.1016/j.jcvp.2024.100183.

30. Nakawesi JS, Wobudeya E, Ndeezi G, Mworozi EA, Tumwine JK. Prevalence and factors associated with rotavirus infection among children admitted with acute diarrhea in Uganda. BMC Pediatr. 2010;10(1):69. doi: 10.1186/1471-2431-10-69.

31. Prabakaran J J, Araya M, Ghebrihiwet A, Andom M, Teklemariam Y, Yosief S, et al. Incidence of Rotavirus Infection and Associated Risk Factors among Children Under 5 Years in Eritrea. Int J Health Sci Res. 2018;8(4):45–54.

32. Martinez-Gutierrez M, Arcila-Quiceno V, Trejos-Suarez J, Ruiz-Saenz J. Prevalence and molecular typing of rotavirus in children with acute diarrhoea in Northeastern Colombia. Rev Inst Med Trop Sao Paulo. 2019;61:e34. doi: 10.1590/S1678-9946201961034.

33. Fan Q. A Clinical Nursing Care Study on the Prevalence of Rotavirus Infection and Acute Diarrhea in Vaccinated Chinese Pediatric Population from 2019–2022. Infect Drug Resist. 2022;15:6129–6142. doi: 10.2147/IDR.S383979.

34. Mahamba D, Hokororo A, Mashuda F, Msanga DR, Bendera EC, Kwiyolecha EN, et al. Prevalence and factors associated with rotavirus infection among vaccinated children hospitalized for acute diarrhea in Mwanza City, Tanzania: a cross sectional study. Open J Pediatr. 2020;10(03):392–403. doi: 10.4236/ojped.2020.103040.

35. Sisay MM. Risk Factors of Rotavirus Outbreak Among Children in Kurmuk District, Benishangul Gumuz Regional State, Ethiopia. Juniper Online J Case Stud. 2018;8(1). doi: 10.19080/JOJCS.2018.08.555730.

36. Salim H, Karyana IPG, Sanjaya-Putra IGN, Budiarsa S, Soenarto Y. Risk factors of rotavirus diarrhea in hospitalized children in Sanglah Hospital, Denpasar: a prospective cohort study. BMC Gastroenterol. 2014;14(1):54. doi: 10.1186/1471-230X-14-54.

